# The Unexpected Protective Role of Thrombosis in Sepsis-Induced Inflammatory Lung Injury Via Endothelial Alox15

**DOI:** 10.1101/2023.03.29.23287934

**Authors:** Colin E. Evans, Xianming Zhang, Narsa Machireddy, You-Yang Zhao

**Affiliations:** Program for Lung and Vascular Biology; and Regeneration Research, Stanley Manne Children’s Research Institute, Ann & Robert H. Lurie Children’s Hospital of Chicago, Chicago, Illinois, USA; Section for Injury Repair and Regeneration Research, Stanley Manne Children’s Research Institute, Ann & Robert H. Lurie Children’s Hospital of Chicago, Chicago, Illinois, USA; Department of Pediatrics, Division of Critical Care, Northwestern University Feinberg School of Medicine, Chicago, Illinois, USA; Department of Pharmacology; School of Medicine, Chicago, Illinois, USA; Department of Medicine, Division of Pulmonary and Critical Care Medicine; School of Medicine, Chicago, Illinois, USA; Feinberg Cardiovascular and Renal Research Institute; Northwestern University Feinberg School of Medicine, Chicago, Illinois, USA

**Keywords:** Acute lung injury, acute respiratory distress syndrome, ALOX15, endothelial cells, inflammation, sepsis, thrombosis

## Abstract

**Background:** Patients with sepsis-induced acute lung injury (ALI)/acute respiratory distress syndrome (ARDS) commonly suffer from severe pulmonary thrombosis, but clinical trials of anti-coagulant therapies in sepsis and ARDS patients have failed. ARDS patients with thrombocytopenia also exhibit increased mortality, and widespread pulmonary thrombosis is often seen in coronavirus disease 2019 (COVID-19) ARDS patients.

**Methods:** Employing different amounts of microbeads to induce various levels of pulmonary thrombosis. Acute lung injury was induced by either lipopolysaccharide i.p. or cecal ligation and puncture. Endothelial cell (EC)-targeted nanoparticle coupled with CDH5 promoter was employed to delivery plasmid DNA expressing the CRISPR/Cas9 system for EC-specific gene knockout or expressing Alox15 for EC-specific overexpression. Additionally, thrombocytopenia was induced by genetic depletion of platelets using *DTR^Pf4Cre^* mice by breeding *Pf4*Cre mice into the genetic background of *DTR* mice.

**Results:** We show that while severe pulmonary thrombosis or thrombocytopenia augments sepsis-induced ALI, the induction of mild pulmonary thrombosis conversely reduces endothelial cell (EC) apoptosis, ALI, and mortality via sustained expression of endothelial arachidonate 15-lipoxygenase (Alox15). Endothelial *Alox15* knockout via EC-targeted nanoparticle delivery of CRISPR/Cas9 plasmid DNA in adult mice abolished the protective impact of mild lung thrombosis. Conversely, overexpression of endothelial *Alox15* inhibited the increases in ALI caused by severe pulmonary thrombosis. The clinical relevance of the findings was validated by the observation of reduced ALOX15-expressing ECs in lung autopsy samples of ARDS patients. Additionally, restoration of pulmonary thrombosis in thrombocytopenic mice also normalized endotoxemia-induced ALI.

**Conclusion:** We have demonstrated that moderate levels of thrombosis protect against sepsis-induced inflammatory lung injury via endothelial Alox15. Overexpression of Alox5 inhibits severe pulmonary thrombosis-induced increase of ALI. Thus, activation of ALOX15 signaling represents a promising therapeutic strategy for treatment of ARDS, especially in sub-populations of patients with thrombocytopenia and/or severe pulmonary thrombosis.

## Introduction

Sepsis is an infection-induced uncontrolled inflammatory response that often results in acute lung injury (ALI)/acute respiratory distress syndrome (ARDS) ^1–3^. Despite improvements in supportive care for patients with ALI/ARDS, including mechanical ventilation and antibiotic therapy, there is currently no effective treatment for ALI/ARDS, and mortality rates for patients with ARDS are as high as 40% ^1–3^. The lack of effective treatments for ALI/ARDS is likely due to an incomplete understanding of the mechanisms that control the pathogenesis of ALI/ARDS. Given that anti-inflammatory therapies have been unsuccessful in the treatment of ARDS ^4, 5^, it is likely that ALI/ARDS is regulated by factors other than inflammation alone. For instance, ALI/ARDS is characterized by increased levels of thrombosis ^1, 6, 7^. Widespread pulmonary thrombosis is often seen in coronavirus disease 2019 (COVID-19) ARDS patients ^8–11^. Thrombocytopenia is also associated with increased mortality in patients with sepsis and ARDS ^12–14^. Increased microvascular thrombosis and thrombocytopenia are hallmarks of disseminated intravascular coagulation (DIC), which is common in sepsis and ARDS patients ^15–17^. However, the impact of thrombosis and thrombocytopenia on ALI is ambiguous ^18^. For example, pulmonary thrombosis impedes the delivery of oxygen and nutrients from blood to the lung, while simultaneously acting as a barrier to prevent bacterial exchange ^19, 20^. Crucially, comprehensive investigations of the impact of varying levels of pulmonary thrombosis and of the underlying mechanisms on ALI/ARDS are absent to date.

Another characteristic of ALI/ARDS is increased lung endothelial cell (EC) apoptosis and injury ^21–23^. A viable lung EC monolayer is essential for the maintenance of vascular homeostasis ^24–26^, while reduced lung EC survival leads to impaired endothelial barrier function, including increased capillary permeability and pulmonary fluid imbalance ^2, 23, 25, 26^. Pulmonary EC apoptosis has been shown in experimental models of sepsis and ALI/ARDS ^27–31^ and in ARDS clinical samples ^21^. Despite these observations, and the possibility that improving lung EC survival could be a useful strategy to combat ALI/ARDS ^32, 33^, the effect of differing severities of lung thrombosis on pulmonary EC survival in ALI/ARDS is also unknown.

Given that anti-coagulants have failed in clinical trials of sepsis/ARDS ^34–37^ and that experimental thrombosis stimulates an angiogenic response ^38, 39^, we hypothesized that mild levels of pulmonary thrombosis could protect against sepsis-induced ALI. In this study, we aimed to determine the impact of varying levels of pulmonary thrombosis on sepsis-induced ALI, and to delineate the underlying molecular mechanisms. We found that the induction of severe levels of pulmonary thrombosis augmented EC apoptosis and ALI in sepsis mice, while a mild level of pulmonary thrombosis conversely protected against ALI and mortality via endothelial arachidonate 15-lipoxygenase (Alox15). EC-targeted overexpression of Alox15 inhibited the increases in ALI caused by severe pulmonary thrombosis. Restoration of mild levels of pulmonary thrombosis inhibited thrombocytopenia-induced increases in ALI and mortality in platelet-depleted sepsis mice. The clinical relevance of the study was supported by our observation that there are markedly reduced ALOX15-expressing ECs in the pulmonary vasculature of ARDS lungs compared with normal lungs.

## Results

### Mild pulmonary thrombosis attenuates, whereas severe thrombosis augments, inflammatory lung injury in endotoxemia mice

Pulmonary thrombosis may have protective or detrimental effects on ALI, so we first investigated the impact of different levels of pulmonary thrombosis on ALI. Wild type (WT) mice received various amounts of polystyrene microbeads (MBs) (i.v.) or vehicle to quantitatively induce pulmonary thrombosis ^39^. At 24hrs later, mice received i.p. lipopolysaccharide (LPS) to induce endotoxemia ^29, 40^, and lung injury was assessed at 36hrs post-LPS (i.e., at the time of peak injury). A dose of 1,000 MBs/mouse led to the greatest reduction in lung vascular permeability (as measured by Evans blue-conjugated albumin [EBA] flux) and neutrophil sequestration (indicated by myeloperoxidase [MPO] activity) ^29, 40^ versus MB-free endotoxic mice (**Fig. 1, A** and **B**). Conversely, increasing the MB number resulted in dose-dependent increases in lung vascular permeability, with 200,000 MBs/mouse resulting in highest levels of lung vascular permeability versus vehicle-treated endotoxic controls (**Fig. 1A**). MPO activity was also markedly increased in mice who received 200,000 MBs compared to MB-free endotoxic controls (**Fig. 1B**). The protective dose of 1,000 MBs/mouse (referred to herein as innocuous) also reduced apoptosis in pulmonary ECs compared with that found in MB-free endotoxic mice, while the detrimental dose of 200,000 MBs/mouse (referred to herein as severe) worsened pulmonary EC apoptosis (**Fig. 1, C** to **E**). In confirmatory studies, we verified that the innocuous MB dose did indeed increase the levels of lung thrombosis above those found in vehicle-treated endotoxic controls, and that this level was further enhanced by the severe dose of MBs (**Fig. 1, F** to **H**).

**Fig. 1.**
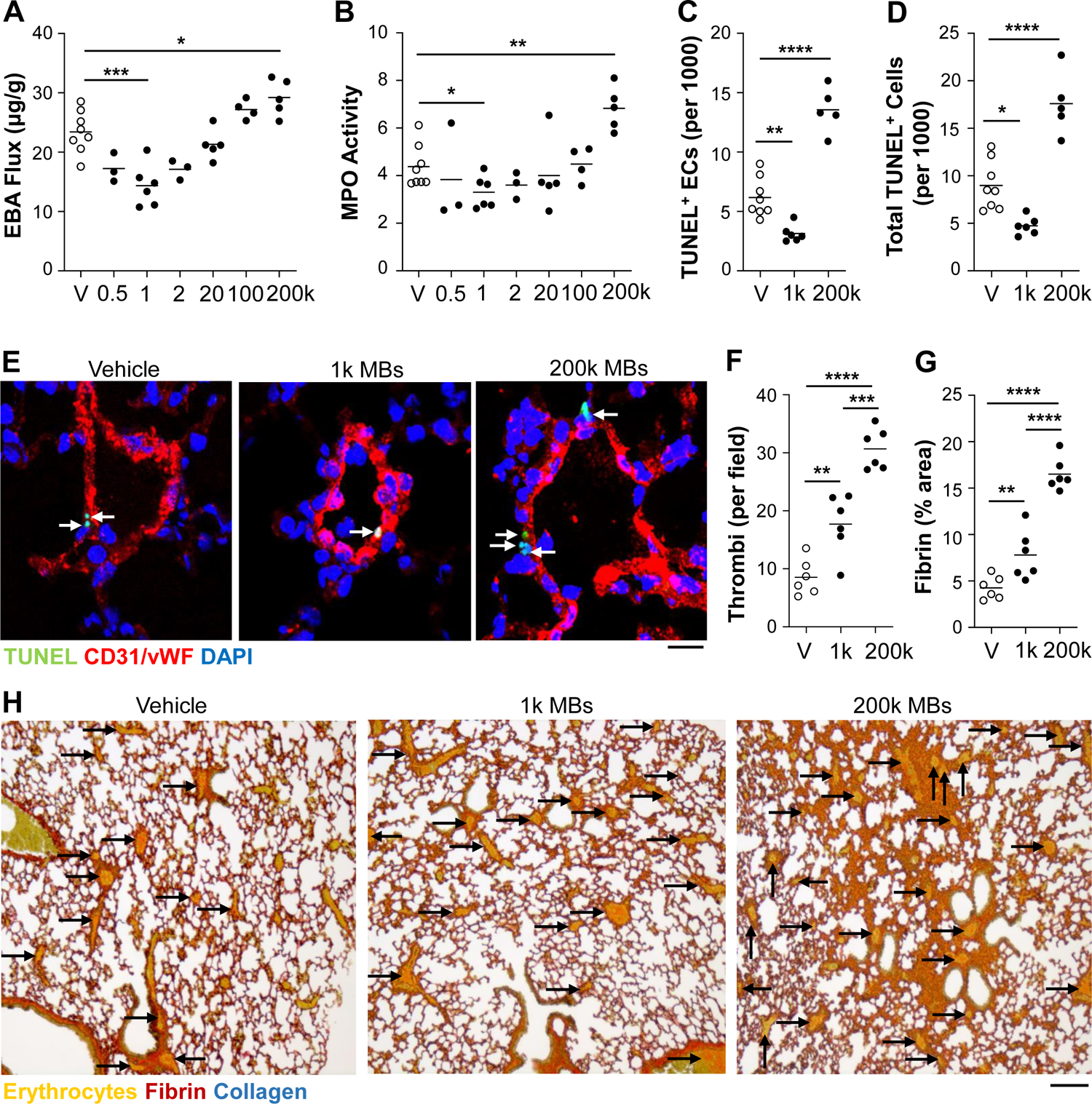
Opposite effects of mild versus severe lung thrombosis on LPS-induced ALI. (**A**) Mice received either PBS vehicle (V) or indicated dose of MBs (i.v.) and 24hrs later received LPS (3mg/kg, i.p.). Lung tissues were collected for analyses at 36hrs post-LPS. EBA flux assay was used to demonstrate that a dose of 1,000 MBs/mouse protected against LPS-induced pulmonary vascular permeability, but higher doses (e.g., 200,000 MBs/mouse) augmented LPS-induced pulmonary vascular permeability. (**B**) MPO activity assay showing that 1,000 MBs/mouse protected against whereas 200,000 MBs/mouse augmented LPS-induced lung neutrophil sequestration. (**C-E**) TUNEL staining and quantification of apoptosis (green). ECs were stained with anti-CD31 and anti-vWF antibodies (ECs, red). Nuclei were counterstained with DAPI (blue). Arrows point to TUNEL^+^ ECs. Scale bar = 10μm. (**F-H**) MSB staining and quantification of thrombi and fibrin area. Arrows indicate fibrin thrombi. Scale bar = 100μm. *P<0.05, **P<0.01, ***P<0.001, and ****P<0.0001; one-way ANOVA with Bonferroni post-tests (**A-D**, **F, G**).

We next extended our studies of the protective effects of the innocuous level of pulmonary thrombosis on lung injury. Consistent with reduced pulmonary vascular permeability (**Fig. 2A**), 1,000 MBs treatment decreased lung edema assessed by lung wet/dry weight ratio in sepsis-challenged but not basal mice (**Fig. 2B**). Assessment of lung histology also revealed a marked reduction in alveolar edema and septal thickening in 1,000 MBs-treated endotoxemic mice compared to vehicle-treated endotoxemic mice (**Fig. 2, C** to **E**).

**Fig. 2.**
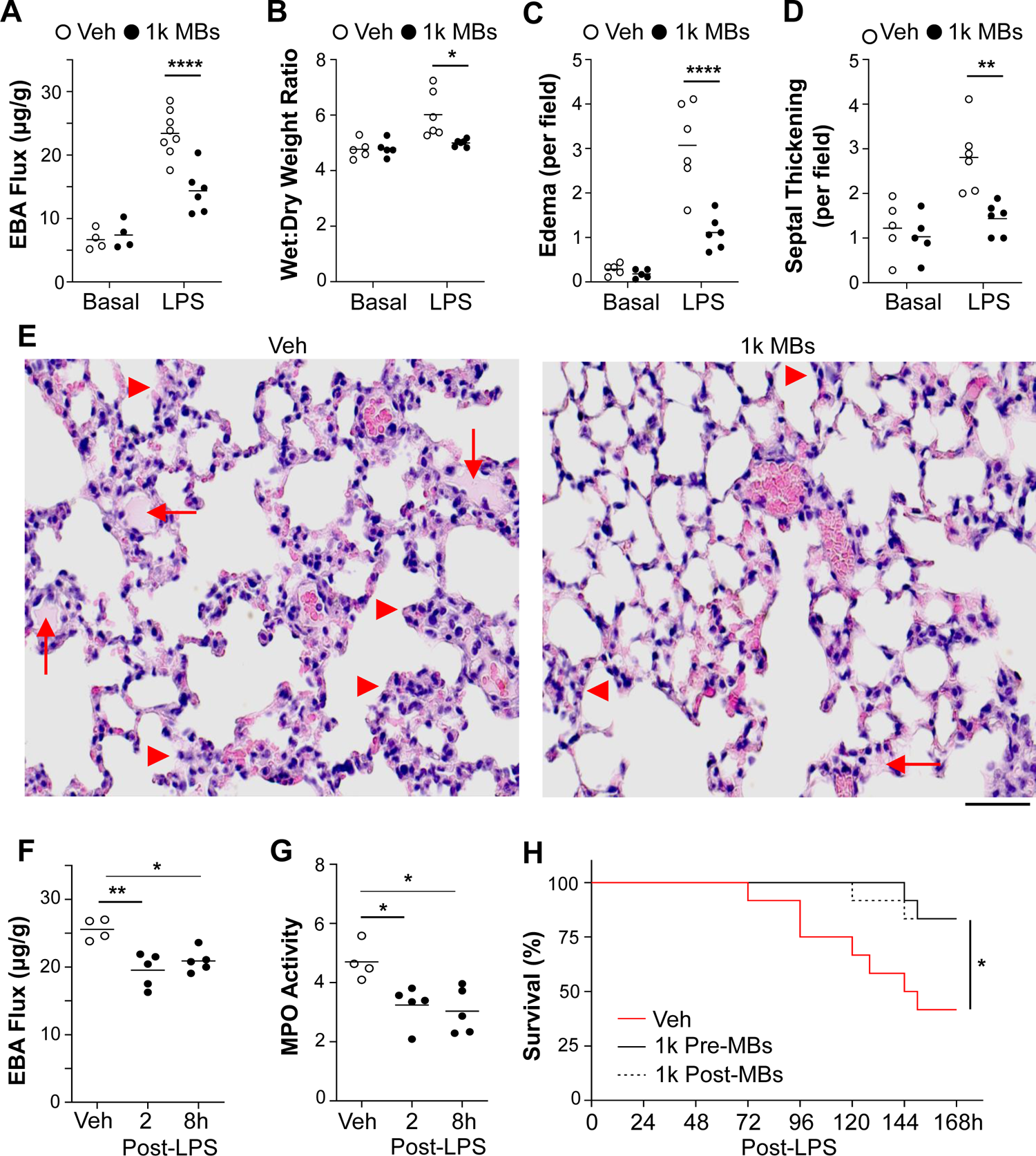
Mild lung thrombosis protects against LPS-induced ALI and improves survival. Mice received PBS (Veh) or 1,000 MBs/mouse (i.v.) and 24hrs later received LPS (3mg/kg, i.p.) or PBS (Basal). Lung tissues were collected for analyses at 36hrs post-LPS. (**A**) EBA flux assay showing that treatment with 1,000 MBs/mouse reduced lung vascular permeability in LPS-treated but not basal mice. Values for 36hrs post-LPS are repeated in Fig 1A and 2A. (**B**) The effect of 1,000 MBs/mouse on basal and LPS-induced lung edema was measured by lung wet/dry weight ratio. Treatment with 1,000 MBs/mouse reduced lung edema in LPS-treated mice. (**C-E**) Histological analysis of H&E-stained lung sections (**E**) showing that 1,000 MBs treatment reduced alveolar edema (**C**), and alveolar septum thickening (**D**) in LPS-treated mice compared to MB-free (Veh) LPS-treated mice. In the representative images (**E**), arrows point to alveolar edema and arrowheads indicate thickened septum. Scale bar = 50µm. (**F, G**) The 1,000 MBs treatment given after LPS challenge was also protective. At 2 or 8hrs after LPS (3mg/kg, i.p.), mice received PBS (Veh) or 1,000 MBs (i.v.). At 36hrs post-LPS, lung tissues were collected for EBA flux assay (**F**) and MPO activity measurement (**G**). (**H**) Both pre- and post-LPS treatments with 1,000 MBs/mouse promoted survival. Mice received 1,000 MBs (i.v.) either 24hrs before (1k Pre-MBs) or 2hrs after (1k Post-MBs) a lethal dose of LPS (6mg/kg, i.p.). Survival was monitored for 1wk. n = 12/group. *P<0.05, **P<0.01, and ****P<0.0001; one-way (**F**, **G**) or two-way ANOVA (**A-D**) with Bonferroni post-tests; log-rank (Mantel-Cox) test (**H**).

To determine whether the innocuous dose of MBs could inhibit inflammatory lung injury when given after sepsis challenge, the 1,000 MBs were administered to WT mice at 2 or 8hrs after LPS challenge. As shown in **Fig. 2, F** and **G**, the protective impact of this innocuous level of pulmonary thrombosis on inflammatory lung injury, evidenced by marked reductions of lung EBA flux and MPO activity at 36hrs post-LPS, was maintained even when the MBs were given at 8hrs post-LPS challenge. Furthermore, treatment with 1,000 MBs promoted survival in mice challenged with a lethal dose of LPS, regardless of whether the MBs were administered before or after sepsis challenge (**Fig. 2H**). Together, these data demonstrate that while severe pulmonary thrombosis enhance inflammatory lung injury, mild pulmonary thrombosis protects against inflammatory lung injury induced by endotoxemia.

### Molecular profiling of genes involved in the protective effects of mild lung thrombosis

To elucidate molecular pathways through which mild pulmonary thrombosis protects against inflammatory lung injury, we next performed RNA sequencing analysis. In these studies, whole lung samples were analyzed from sepsis-free WT mice (basal group) and from LPS-treated WT mice without MBs (vehicle group) or with mild (1,000 MBs/mouse) or severe (200,000 MBs/mouse) pulmonary thrombosis. Lung samples were collected for analysis at 24hrs post-LPS. Of the 15,531 genes detected by RNA sequencing analysis, we selected the genes whose expression levels varied from one group to another by ≥ 2-fold, yielding a list of 82 genes. Of these 82 genes, 12 are known to regulate cell survival (**Fig. 3A**). Of these 12 genes, the expression patterns of 9 genes were validated in whole lung samples by qRT-PCR analysis (**Fig. 3B** and **Fig. S1**). The expression of 6 of these genes was increased in lung samples of LPS mice treated with 1,000 MBs compared with mice receiving LPS alone, but not in LPS mice treated with 200,000 MBs (**Fig. 3B**). Quantitative (Q)RT-PCR analysis of isolated cells further showed that these 6 genes were increased in either pulmonary non-ECs (i.e., Ghrl and Ms4a1), pulmonary ECs (i.e., Alox15 and Cd22), or both pulmonary non-ECs and ECs (i.e., Pla2g2d and Serpinb2) of 1,000 MBs-treated mice (**Fig. 3, C** and **D**). These data suggested that mild pulmonary thrombosis could protect against inflammatory lung injury via attenuation of sepsis-induced decreases of the expression of factors that regulate cell survival.

**Fig. 3.**
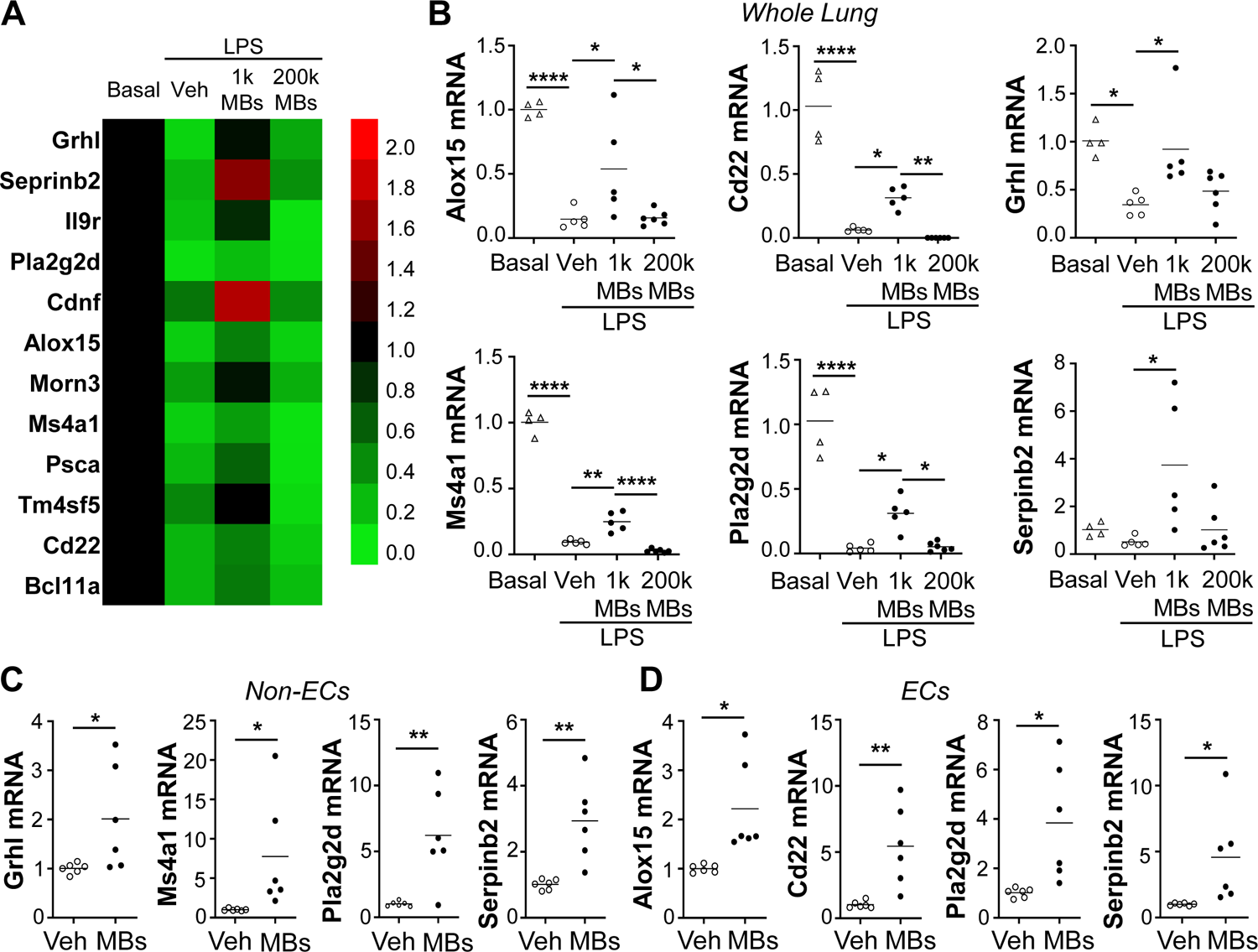
The effects of lung thrombosis on expression of survival factors in lungs of sepsis-challenged mice. Mice received PBS (Veh) or indicated dose of MBs (i.v.) and 24hrs later were challenged with LPS (3mg/kg, i.p.). At 24hrs post-LPS, lung tissues were collected for RNA sequencing and qRT-PCR analyses. (**A**) Expression heatmap of survival genes that were identified by RNA sequencing of whole lung samples to be downregulated in vehicle-treated LPS mouse lungs versus basal mice and restored in LPS mice receiving 1,000 but not 200,000 MBs. (B) QRT-PCR analysis confirming the effect of varying extents of lung thrombosis on the expression patterns of survival genes in whole lungs of basal compared with sepsis-challenged mice. (**C, D**) QRT-PCR analysis of expression of the survival factors in isolated pulmonary cells from LPS-challenged mice, identifying gene induction by 1,000 MBs (MBs) in non-ECs (CD31^-^ cells) (**C**) and/or ECs (CD31^+^ cells) (**D**). *Alox15* and *Cd22* were the 2 genes that were upregulated only in ECs. *P<0.05, **P<0.01, and ****P<0.001; one-way ANOVA with Bonferroni post-test (**B**) or independent t-tests (**C**, **D**).

### Endothelial Alox15 mediates the protective effects of mild lung thrombosis

Given that mild lung thrombosis resulted in increased expression of Alox15 and Cd22 in pulmonary ECs but not non-ECs, we next determined whether either of these factors was responsible for the protective effects of mild thrombosis. We employed nanoparticle-targeted delivery of an all-in-one CRISPR plasmid DNA expressing Cas9 under control of the human *CDH5* promoter and guide RNA by the *U6* promoter, to delete either of these 2 genes selectively in ECs of adult mice (**Fig. 4A**). Consistent with a previous study, which demonstrated ∼80% decreases in protein expression ^41^, qPCR analysis demonstrated an approximately 40% genome editing efficiency in the *Alox15* or *Cd22* locus in ECs but not non-ECs of the respective CRISPR plasmid-transduced mice (**Fig. S2, A** and **B**). We confirmed an 80% decrease of EC-specific Alox15 at the protein level (**Fig. 4B**). Seven days after nanoparticle:CRISPR plasmid DNA administration, the mice receiving plasmid DNA expressing the scrambled guide RNA (WT group) or genome-edited mice were administered either vehicle or 1,000 MBs followed by LPS challenge at 24hrs after vehicle or MBs, and lungs were assessed at 36hrs post-LPS (**Fig. 4A**). As shown in **Fig. 4C**, CRISPR-mediated disruption of endothelial *Alox15* but not *Cd22* reversed the 1,000 MBs-induced decrease in pulmonary EBA flux. Consistently, lung MPO activity in endothelial *Alox15*-but not *Cd22*-genome-edited mice treated with 1,000 MBs was markedly elevated compared to 1,000 MBs-treated WT mice (**Fig. 4D**). These data provide clear evidence that mild lung thrombosis protects against inflammatory lung injury via endothelial Alox15 but not Cd22. Furthermore, we observed endothelial-specific disruption of *Alox15* restored histological lung injury in 1,000 MBs-treated mice to levels comparable with vehicle-treated WT mice, in contrast to 1,000 MBs-treated WT mice (**Fig. 4, E** and **F**). Next, we showed that the inhibition of endothelial apoptosis by 1,000 MBs in WT mice was also reversed in *Alox15* genome-edited mice (**Fig. 4, G** to **I**). These data together demonstrate that mild lung thrombosis reduces inflammatory lung injury via endothelial Alox15.

**Fig. 4.**
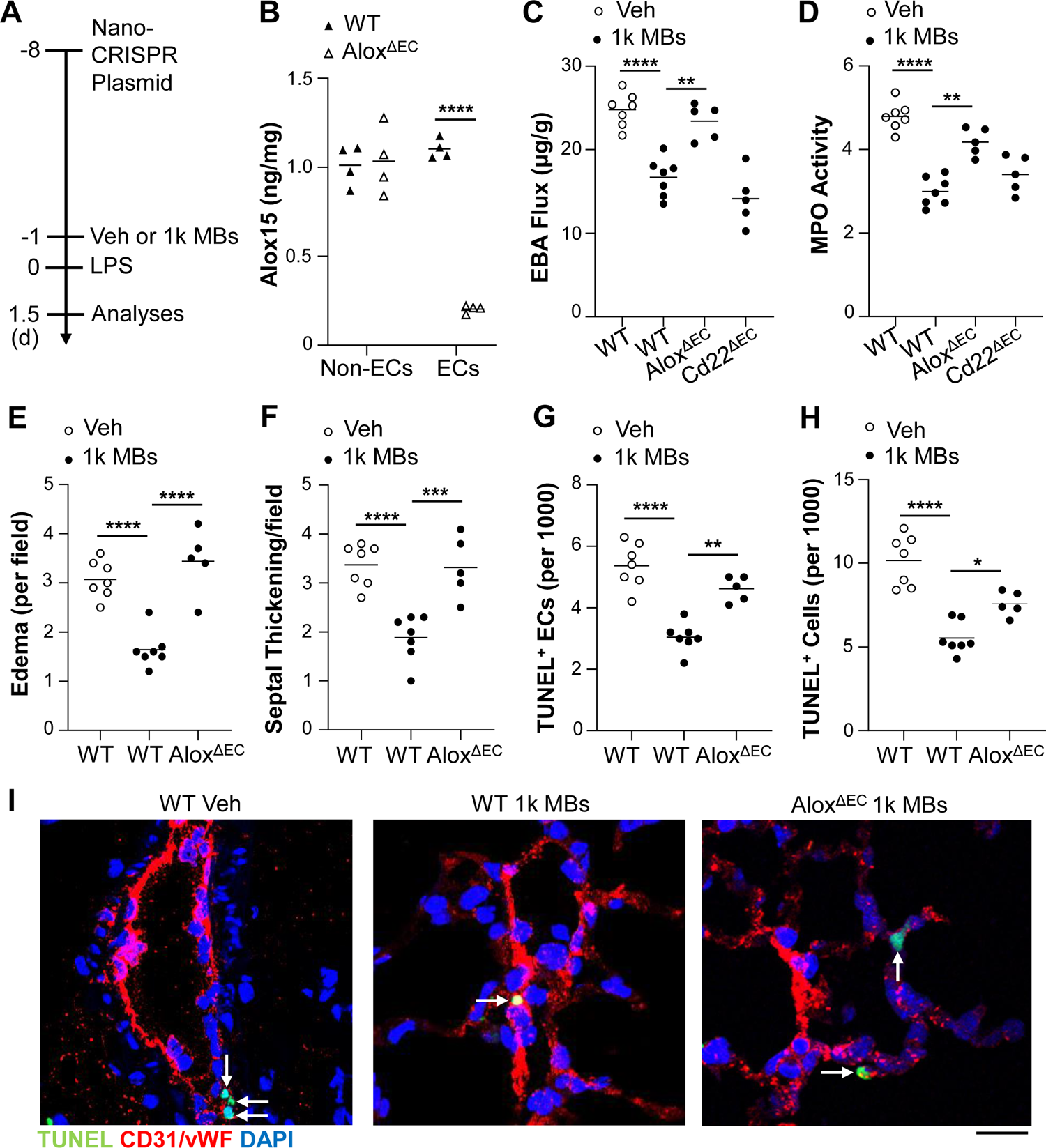
Mild lung thrombosis protects against LPS-induced ALI via endothelial Alox15. (**A**) Diagrammatical representation of the experimental procedure. Adult mice received a mixture of nanoparticle:CRISPR*^CDH5^* plasmid DNA expressing Cas9 under control of the human *CDH5* promoter and gene-specific (*Alox15* or *Cd22*) guide RNA (designated as *Alox^ΔEC^* or *Cd22^ΔEC^*, respectively) or scrambled guide RNA (WT) driven by the *U6* promoter (40ug plasmid DNA/mouse, i.v.). At 7days later, mice received PBS (Veh) or 1,000 MBs (i.v.) followed by LPS challenge (3mg/kg, i.p.) at 24hrs later. Lung tissues were collected at 36hrs post-LPS for various analyses. (**B**) ELISA analysis demonstrating diminished Alox15 expression in lung ECs but not in non-ECs isolated from CRISPR/*Alox15* gRNA-plasmid-administered basal mice. Lung tissues were collected from mice at 7 days post-nanoparticle delivery of plasmid DNA. (**C**) CRISPR plasmid DNA-mediated disruption of endothelial *Alox15* but not endothelial *Cd22* reversed the decrease in pulmonary vascular permeability caused by treatment with 1,000 MBs/mouse. (**D**) Knockdown of endothelial *Alox15* but not *Cd22* also reversed the 1,000 MBs treatment-elicited decrease in MPO activity. (**E, F**) Quantitative analyses of H&E-stained lung sections showing reversal of 1,000 MB treatment-induced decreases in alveolar edema (**E**), and septum thickening (**F**) in *Alox^ΔEC^* mice. (**G-I**) Quantifications of pulmonary EC apoptosis (**G**) and total apoptosis (**H**) in LPS-challenged mice. Lung cryosections were stained with FITC-TUNEL (apoptosis, green) and anti-CD31/-vWF antibodies (ECs, red). Nuclei were counterstained with DAPI (blue). Arrows point to TUNEL^+^ ECs (**I**). Scale bar = 10μm. *P<0.05, **P<0.01, ***P<0.001, and ****P<0.0001; two-way (**B**) or one-way ANOVA with Bonferroni post-tests (**C-H**).

### Overexpression of Alox15 in ECs inhibits the detrimental effect of severe lung thrombosis in septic mice

Given that severe pulmonary thrombosis failed to reverse the sepsis-induced downregulation of Alox15, we next determined whether restoration of expression of *Alox15* in ECs could reverse the deleterious impact of severe lung thrombosis on ALI. Twenty hours after administration of the mixture of nanoparticles:plasmid DNA expressing *Alox15* under the control of *CDH5* promoter or empty vector, the mice were challenged with LPS, then treated with vehicle or 200,000 MBs/mouse at 2hrs after LPS. Lungs were assessed at 36hrs post-LPS (**Fig. 5A**). QRT-PCR analysis demonstrated a marked increase in *Alox15* mRNA expression selectively in lung ECs of *Alox15* plasmid-transduced mice compared with vector mice (**Fig. S3**), which was confirmed at the protein level (**Fig. 5B**). The 200,000 MBs-induced increase in pulmonary vascular permeability was inhibited by EC-specific *Alox15* overexpression in *Alox15* plasmid-transduced mice (**Fig. 5C**). Histological increases in lung edema and septal thickening caused by 200,000 MBs/mouse were also abolished by EC-specific *Alox15* overexpression in *Alox15* plasmid-transduced mice with severe lung thrombosis (**Fig. 5, D** and **E**). Similarly, the severe lung thrombosis-induced increase of lung EC apoptosis was also inhibited by EC-specific *Alox15* overexpression (**Fig. 5, F** to **H**). These data suggest that increases in ALI resulting from severe lung thrombosis can be reversed by an EC-targeted gene therapy approach to restore endothelial Alox15 expression.

**Fig. 5.**
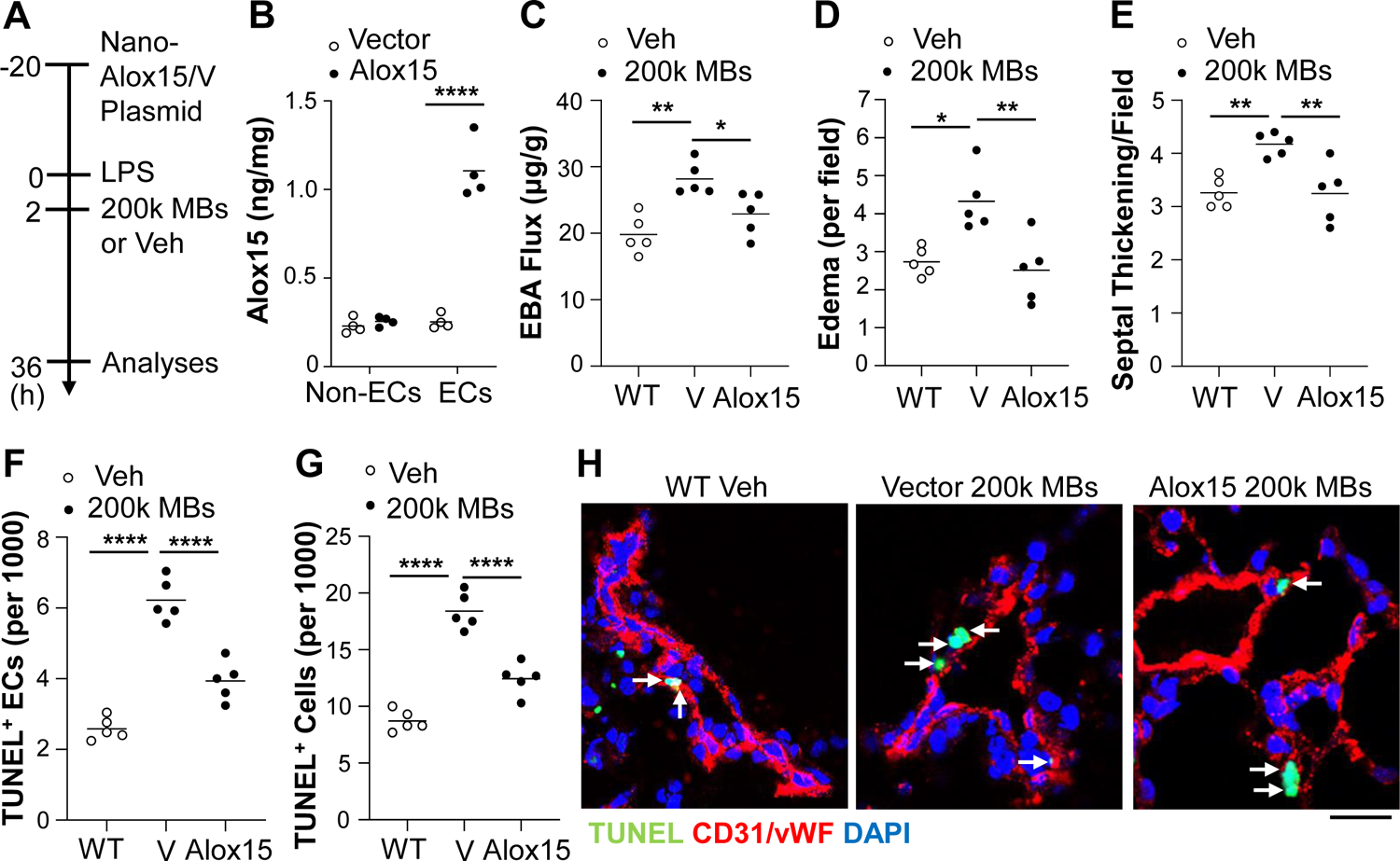
Overexpression of endothelial Alox15 inhibits the detrimental effects of severe lung thrombosis on sepsis-induced lung injury. (**A**) Diagrammatical representation of the experimental procedure. (**B**) ELISA analysis demonstrating a marked increase of Alox15 expression in isolated lung ECs but not non-ECs of *Alox15* plasmid (Alox15)-transduced mice compared to empty vector (V) mice. Human *CDH5* promoter was employed in the plasmid DNA. (C) EBA flux assay demonstrating endothelial *Alox15* overexpression reversed the 200,000 MBs-elicited increase of pulmonary vascular permeability in LPS mice. (**D, E**) Quantification of H&E-stained mouse lung sections showing that overexpression of endothelial *Alox15* inhibited the severe thrombosis-induced increases in alveolar edema (**D**) and septum thickening (**E**). (**F**-**H**) Quantifications of apoptosis in the lung demonstrating that 200,000 MBs/mouse-induced increases in EC apoptosis (**F**) and total apoptosis (**G**) was inhibited by EC-specific *Alox15* overexpression. Lung cryosections were stained with FITC-TUNEL (green) and with anti-CD31 and -vWF antibodies (ECs, red). Nuclei were counterstained with DAPI (blue). In the representative micrographs (**F**), arrows point to TUNEL^+^ ECs. Scale bar = 10μm. *P<0.05, **P<0.01, and ****P<0.0001; one-way ANOVA with Bonferroni post-tests (**B-G**).

### Restoration of mild levels of pulmonary thrombosis attenuate thrombocytopenic ALI

Given that thrombocytopenia is associated with decreased survival in sepsis patients ^12, 13^ and with increased risk of developing ARDS, as well as decreased survival in ARDS patients ^14^, we next wished to determine whether thrombocytopenia-induced increases in ALI could be inhibited through restoration of an innocuous level of lung thrombosis. To generate thrombocytopenic mice, we used a genetic mouse model of platelet depletion in which thrombocytopenia is induced by diphtheria toxin (DT) treatment of mice expressing the DT receptor (DTR) in platelets and megakaryocytes controlled by *Pf4*Cre ^42^. We first showed that circulating levels of platelets are reduced in this *DTR^Pf4Cre^* model (i.e., PF4 group) to <10% of levels found in *DTR^Pf^*^4^*^-^* controls (i.e., WT group); this reduction occurred by day 2 after DT treatment and remained suppressed to this extent for at least 3days (**Fig. S4A**) without altering leukocyte numbers (**Fig. S4, B** to **G**). In subsequent studies of thrombocytopenic mice, LPS was administered at the start of this period of platelet depletion and lungs were assessed at the time point of peak injury (i.e., 36hrs post-LPS). While thrombocytopenia did not alter basal levels of lung thrombosis in LPS-free mice, sepsis challenge gave rise to increases in lung thrombosis in WT mice, and these increases were attenuated by thrombocytopenia (**Fig. S5. A** to **C**). Concomitantly, in LPS-treated mice, thrombocytopenia resulted in exacerbated levels of inflammatory lung injury, evidenced by increased lung EBA flux (**Fig. S6A**) and MPO activity (**Fig. S6B**), as well as histological evidence of increased lung edema and alveolar septum thickening (**Fig. S6, C** to **E**). These data demonstrate that thrombocytopenia impairs sepsis-induced lung thrombosis and increases inflammatory lung injury.

To determine whether thrombocytopenia-induced increases in inflammatory lung injury following sepsis challenge could be rescued by restoration of an innocuous level of lung thrombosis, we next administered i.v. MBs at 24hrs prior to LPS challenge in thrombocytopenic mice (**Fig. 6A**). Starting with the protective dose of 1,000 MBs/mouse identified above in WT mice, we found that increasing doses of i.v. MBs gave rise to increasing levels of lung thrombosis in thrombocytopenic sepsis-challenged mice, and that these increases occurred in a dose-dependent manner (**Fig. 6, B** and **C**). The dose of MBs that restored lung thrombosis in PF4 mice to the levels found in sepsis-challenged WT mice was 2,000 MBs/mouse (**Fig. 6, B** and **C**). Importantly, 2,000 MBs/mouse did not alter the basal levels of lung EBA flux and MPO activity in either platelet-replete (i.e., WT) or thrombocytopenic mice (data not shown). We then assessed inflammatory lung injury in LPS-treated mice and found that this dose of 2,000 MBs/mouse completely abolished the thrombocytopenia-induced increases in inflammatory lung injury as assessed by histology (**Fig. 6, D** to **F**), EBA flux (**Fig. 6G**), and MPO activity (**Fig. 6H**). A dose of 2,000 MBs/mouse elicited the optimal protective effect compared to 1,000 MBs/mouse, 3,000 MBs/mouse, and 5,000 MBs/mouse (**Fig. 6, G** and **H**). However, a high dose of 100,000 MBs/mouse, resulting in excessive thrombosis (**Fig. 6C**), had no protective effects in PF4 thrombocytopenic mice and returned the levels of sepsis-induced inflammatory lung injury to that found in MB-free thrombocytopenic mice (**Fig. 6, D** to **F**).

**Fig. 6.**
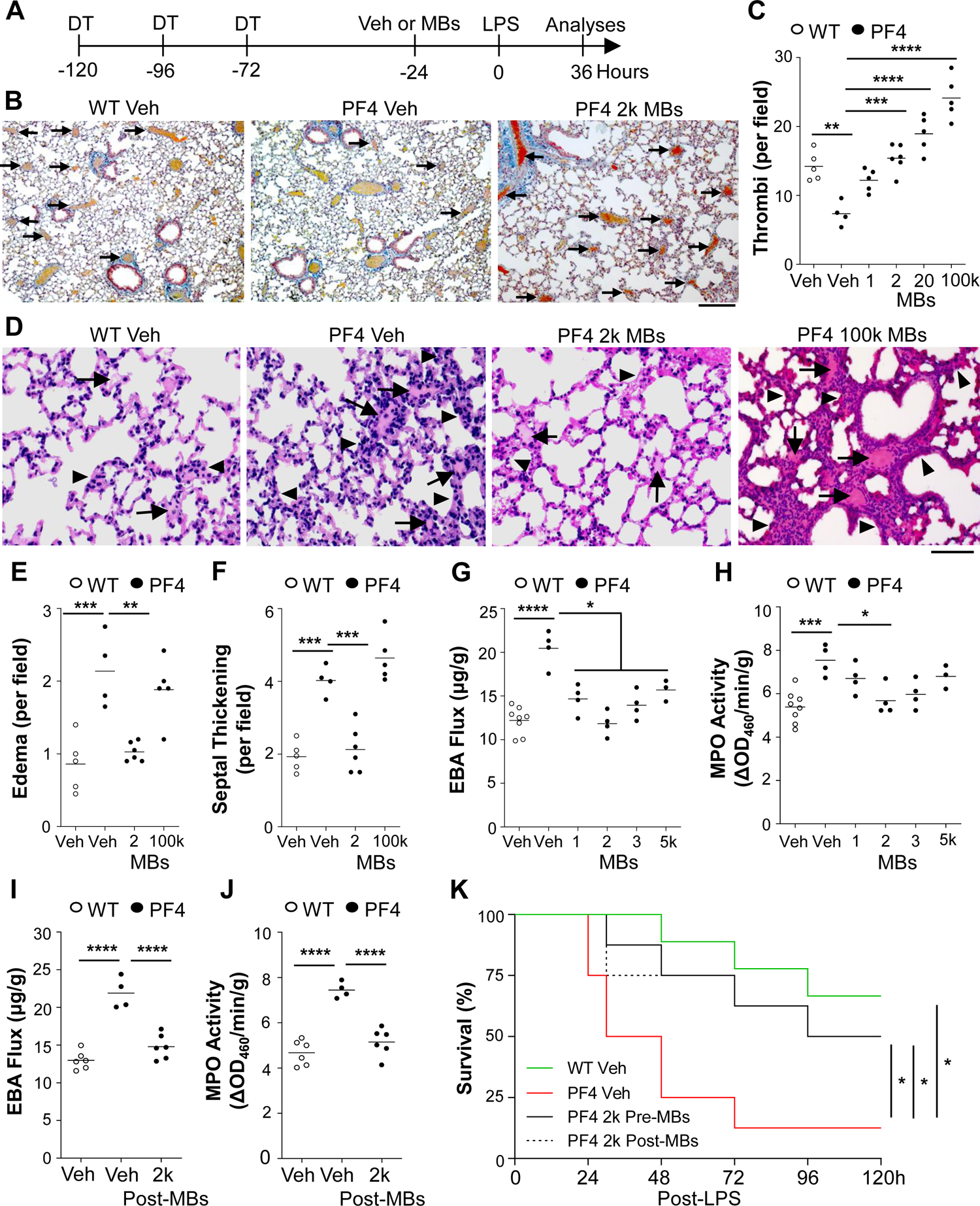
Restoration of mild pulmonary thrombosis inhibits thrombocytopenia-enhanced inflammatory lung injury in sepsis-challenged mice. (**A**) Diagrammatical representation of the experimental procedure. *DTR^Pf4Cre^* (PF4) mice or *DTR^Pf4-^* (WT) mice received DT (20ng/g/day, i.p.) then PBS (Veh) or indicated dose of MBs (i.v.) followed by LPS (3mg/kg, i.p.). (**B**) Representative images of MSB-stained lung cross-sections, showing apparent restoration of thrombosis in thrombocytopenia PF4 mice by 2,000 MB treatment. Arrows indicate fibrin thrombi. Scale bar = 100μm. (**C**) Quantifications of thrombi number revealing that thrombocytopenia resulted in reduced pulmonary thrombosis in sepsis-challenged mice, which was reversible by MB treatment. (**D**) Representative images of H&E lung cross-sections. Arrows indicate alveolar edema. Arrowheads point to septal thickening. Scale bar = 50μm. (**E, F**) Quantifications of alveolar edema and septal thickening. (**G**) EBA flux assay demonstrating that thrombocytopenia-induced increase of pulmonary vascular permeability in PF4 mice was inhibited by treatment with low doses (1,000-5,000) of MBs/mouse. (**H**) MPO activity assay. (**I**) Thrombocytopenia-induced increase of pulmonary vascular permeability in PF4 LPS mice was reversed by post-sepsis restoration of lung thrombosis. 2,000 MBs or PBS (Veh) were given at 2hrs after LPS. (**J**) MPO activity. (**K**) Treatment with 2,000 MBs improved survival in thrombocytopenic LPS-challenged mice. At 48hrs after final DT injection, mice received 4mg/kg LPS (i.p.). At 24hrs before (Pre-MBs) or 2hrs after (Post-MBs) LPS challenge, mice received PBS (Veh) or 2,000 MBs. n = 8-9/group. *P<0.05, **P<0.01, ***P<0.001, and ****P<0.001; one-way ANOVA with Bonferroni post-test (**C**, **E-J**); log-rank (Mantel-Cox) test (**K**).

Next, we observed that the beneficial dose of 2,000 MBs/mouse inhibited thrombocytopenia-induced increases in lung EBA flux and MPO activity, even when the MBs were given at 2hrs after LPS (**Fig. 6, I** and **J**). Consistent with these findings, thrombocytopenia reduced survival in PF4 mice challenged with lethal doses of LPS, but survival was improved by the restoration of an innocuous level of lung thrombosis either before or after sepsis challenge (**Fig. 6K**). These data suggest that thrombocytopenia worsens ALI at least in part through impaired lung thrombosis, and that restoration of an innocuous level of lung thrombosis can protect against thrombocytopenia-induced ALI.

### Mild pulmonary thrombosis is also protective against inflammatory lung injury induced by polymicrobial sepsis via endothelial Alox15

We next determined the effects of various levels of pulmonary thrombosis on inflammatory lung injury in a mouse model of polymicrobial sepsis induced by cecal ligation and puncture (CLP). Similar to endotoxemia mice, sepsis-induced increases of EBA flux, MPO activity, endothelial apoptosis, and histological lung injury were reduced in mice treated with 1,000 MBs/mouse whereas exaggerated in mice treated with 200,000 MBs/mouse (**Fig. 7, A** and **B**, **Fig. S7, and Fig. S8**). To determine if the protective effects of mild pulmonary thrombosis are also mediated by endothelial Alox 15, we employed EC-targeted nanoparticle delivery of CRISPR/Cas9 plasmid to selectively knockout *Alox15* in ECs in adult mice. The mice were then treated with 1,000 MBs/mouse followed by CLP challenge. As shown in **Fig. 7, C-I**, 1,000 MB-induced decreases in EBA flux, MPO activity, endothelial apoptosis, and histological lung injury in WT mice were abrogated in endothelial *Alox15* knockout mice.

**Fig. 7.**
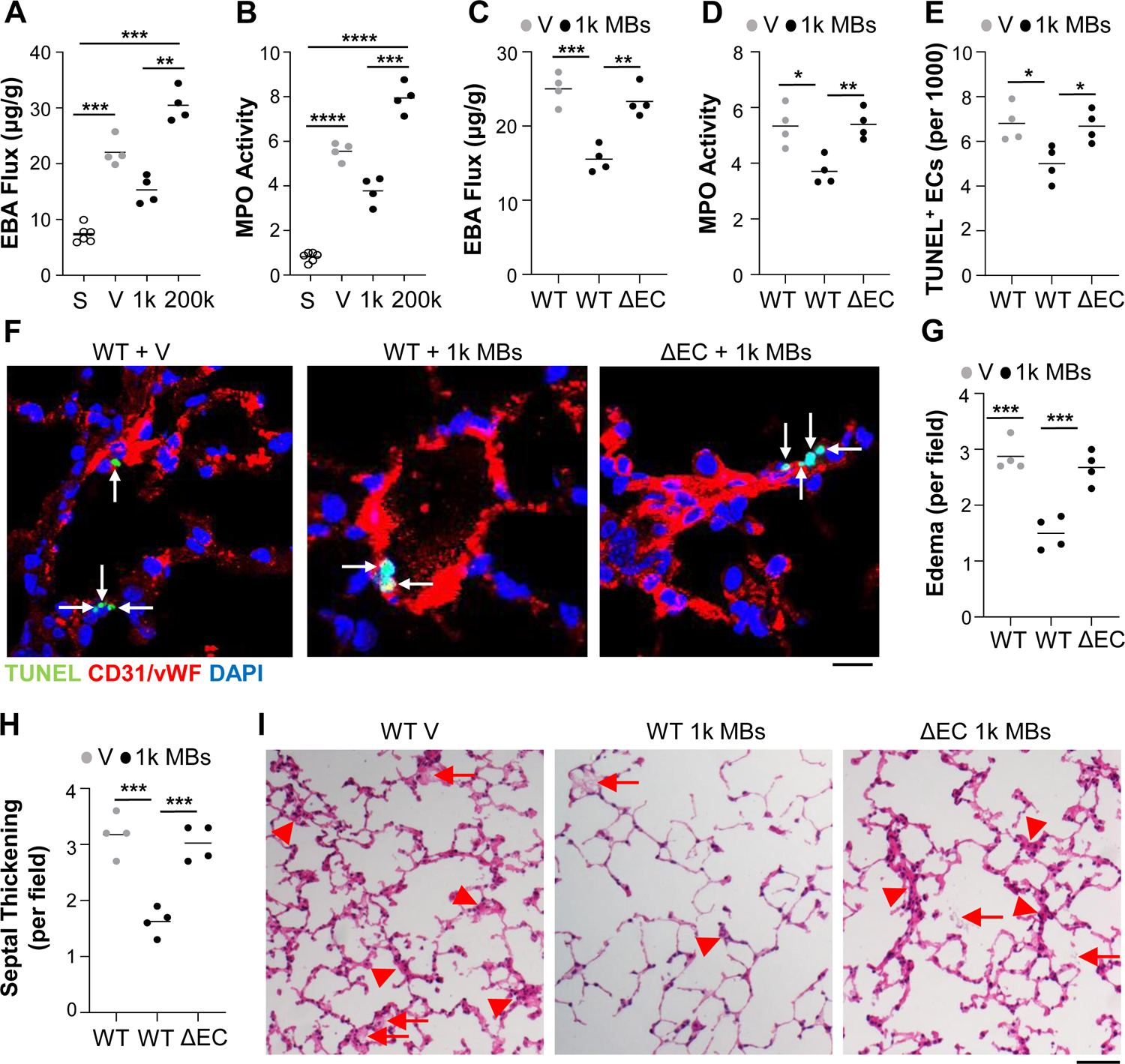
Mild lung thrombosis protects against CLP-induced ALI via endothelial Alox15. (**A**) Mice received either PBS vehicle (V) or indicated dose of MBs (i.v.) and 24hrs later received CLP. Lung tissues were collected for analyses at 36hrs post-CLP. EBA flux assay was used to demonstrate that a dose of 1,000 MBs/mouse protected against CLP-induced pulmonary vascular permeability, but the high dose (i.e., 200,000 MBs/mouse) augmented CLP-induced pulmonary vascular permeability. (**B**) MPO activity assay showing that 1,000 MBs/mouse protected against whereas 200,000 MBs/mouse augmented CLP-induced lung neutrophil sequestration. (**C-I**) Adult mice received a mixture of nanoparticle:CRISPR*^CDH5^* plasmid DNA expressing Cas9 under control of the human *CDH5* promoter and gene-specific (*Alox15*) guide RNA (designated as ΔEC) or scrambled guide RNA (WT) driven by the *U6* promoter. At 7days later, mice received PBS vehicle (V) or 1,000 MBs (i.v.) followed by CLP challenge at 24hrs later. Lung tissues were collected at 36hrs post-CLP for various analyses. CRISPR plasmid DNA-mediated disruption of endothelial *Alox15* reversed the decrease in pulmonary vascular permeability caused by treatment with 1,000 MBs/mouse (**C**). Knockdown of endothelial *Alox15* also reversed the 1,000 MBs treatment-elicited decrease in MPO activity (**D**). TUNEL staining and quantification of apoptosis (green) (**E, F**). ECs were stained with anti-CD31 and anti-vWF antibodies (ECs, red). Nuclei were counterstained with DAPI (blue). Arrows point to TUNEL^+^ ECs. Scale bar = 10μm. Quantitative analyses of H&E-stained lung sections showing reversal of 1,000 MB treatment-induced decreases in alveolar edema (**G**) and septum thickening (**H**) in *Alox^ΔEC^* mice. In the representative images (**I**), arrows point to alveolar edema and arrowheads indicate thickened septum. Scale bar = 50µm. *P<0.05, **P<0.01, ***P<0.001, and ****P<0.0001; one-way ANOVA with Bonferroni post-tests (**A-D, F, G**).

### ALOX15^+^ ECs are diminished in lung samples of ARDS patients

To explore the clinical relevance of our findings, we assessed endothelial *ALOX15* expression in human lungs. *In situ* RNA hybridization (RNAscope) assays of lung sections taken postmortem from 6 ARDS patients (2 males, 4 females, 34-67 years old) and 6 unused normal donor lungs (4 males, 2 females, 43-56 years old) revealed that the proportion of lung ECs that were *ALOX15*^+^ was markedly reduced in ARDS patients compared with normal controls (**Fig. 8, A** and **B**). These data suggest that ALOX15 in human lung ECs is also protective against inflammatory lung injury.

**Fig. 8.**
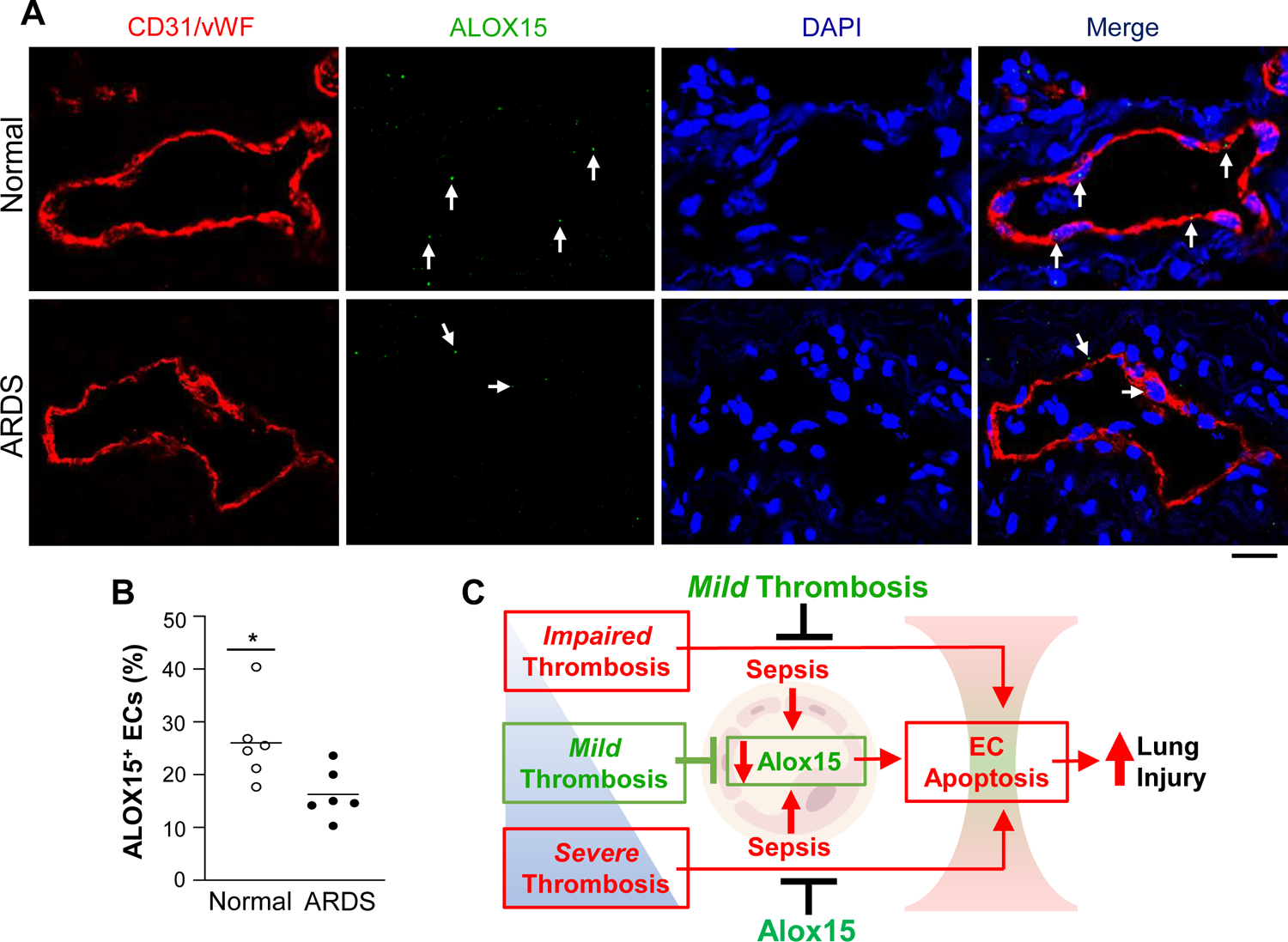
ALOX15^+^ ECs were markedly reduced in lungs of non-survival ARDS patients. (**A**) Representative images of *in situ* RNA hybridization (RNAscope) assays for *ALOX15* expression (green) in archived postmortem lung paraffin sections from ARDS patients and unused donor controls. ECs were immunostained with anti-CD31 and -vWF antibodies (red). Nuclei were counterstained with DAPI (blue). Arrows indicate *ALOX15*^+^ lung ECs. Scale bar = 10μm. (**B**) Quantification of *ALOX*15^+^ ECs in lung sections showing reduced *ALOX15*^+^ ECs in lung sections from ARDS patients compared with normal controls. *P<0.05; unpaired t-test. (**C**) Diagrammatical summary of key findings. Mild lung thrombosis reduces sepsis-induced inflammatory lung injury, whereas severe or impaired lung thrombosis induced by thrombocytopenia augments sepsis-induced inflammatory lung injury. Restoration of lung thrombosis in platelet-depleted mice inhibits the thrombocytopenia-induced increase in lung injury and promotes survival. The protective effects elicited by mild lung thrombosis are mediated by inhibiting the sepsis-induced downregulation of endothelial Alox15, which promotes EC survival.

## Discussion

Our studies demonstrate that while a diminished or severely increased level of lung thrombosis promotes sepsis-induced ALI, a mildlevel of lung thrombosis induces an EC survival response and protects against ALI (**Fig. 8C**). Restoration of a mild level of lung thrombosis in platelet-depleted mice inhibits the thrombocytopenia-induced increases in lung injury and promotes survival. Furthermore, we show in 2 discrete mouse models of sepsis-induced ALI, that the protective effects elicited by mild lung thrombosis are mediated by endothelial Alox15. Loss of endothelial *Alox15* abolished the protective effects, while overexpression of endothelial *Alox15* protected against severe thrombosis-induced increases in EC death and lung injury. In ARDS patients, *ALOX15*^+^ ECs of the pulmonary vasculature were markedly reduced. Together, these studies provide a comprehensive understanding of the role of varying levels of lung thrombosis in the pathogenesis of inflammatory lung injury. Activation of ALOX15 signaling (e.g., via EC-targeted nanoparticle delivery of the *ALOX15* gene) is a potential effective therapeutic strategy for the treatment of ALI/ARDS caused by sepsis, particularly in patient sub-populations with thrombocytopenia or severe pulmonary thrombosis.

In our studies, increasing doses of i.v. MBs were used to quantitatively induce lung thrombosis in a stepwise manner. The MB size used in the current study (15 µm diameter) was selected to ensure seeding into the lung microvasculature ^39^. The same kind of MBs were also used to induce lung thrombosis in a previous study of lung tumorigenesis in WT mice and triggered a pro-angiogenic response in the pulmonary vasculature ^39^. We showed herein that the innocuous level of lung thrombosis did not induce lung injury or inflammation in LPS-free (basal) mice. It should be noted here that MBs have been previously used to induce chronic thromboembolic pulmonary hypertension (CTEPH); the 15 µm MBs used in our study, however, are almost 6 times smaller than the MBs used to induce CTEPH in rats (85µm diameter) ^43^ and 20 times smaller than the MBs in dogs (100-300µm diameter) ^44^. Furthermore, in a previous rodent study of CTEPH, a one-off dose of 388,000 MBs/rat (85µm diameter) failed to induce a hypertensive response at 3 or 6wks after administration ^43^, while repetitive administration of even larger MBs (100-300µm diameter) every 3-4 days takes >100days and >25,000 MBs/animal to induce a hypertensive response in dogs ^44^. Although we did not assess pulmonary hypertension in the current study, the one-off doses of 1,000 or 2,000 MBs/mouse did not cause any discernable behavioral or respiratory changes or unexpected deaths in this study.

It has been suggested that severe levels of lung thrombosis could worsen ALI through ischemia-induced cell death ^18^. Our study shows that a dose of 200,000 MBs/mouse does indeed increase EC apoptosis and enhance ALI. Conversely, mild lung thrombosis induced by 1,000 MBs/mouse protected against ALI in septic WT mice. It has also been suggested that thrombosis could protect against ALI by preventing bacterial transport between the lung and blood, by providing a binding site for pro-survival factors and cytokines, and by blocking leaky endothelial junctions ^18, 45^. Here we show that mild lung thrombosis protects against lung EC apoptosis and ALI and promotes mouse survival via endothelial Alox15. Alox15 is an enzyme producing 12- and 15-hydroxyeicosatetraenoic acid from arachidonic acid. In previous studies ^46–48^, allergen-induced airway inflammation was diminished in global *Alox15* knockout mice, while global *Alox15* deficiency increased inflammation and necroptosis in a murine dermatitis model, and *Alox15*-overexpressing transgenic rabbits were protected against periodontitis. In experimental models of murine ALI, global *Alox15* deletion or systemic inhibition has been previously shown to decrease ALI following aerosolized LPS inhalation or intratracheal hydrochloric acid instillation, which is largely mediated by hematopoietic Alox15, but global *Alox15* deletion instead increased ALI following *A. fumigatus* infection. The apparent incongruence between reported findings could be due to differences in experimental models and the use of global gene targeting methods, as well as disparities resulting from pro-versus anti-inflammatory effects of Alox15 and its enzymatic products ^49^. Nevertheless, we found that *Alox15* expression in total lung and in lung ECs was downregulated following LPS challenge and that the innocuous level of lung thrombosis could attenuate sepsis downregulation of endothelial *Alox15* expression. Nanoparticle delivery of the CRISPR system-mediated disruption of endothelial *Alox15* inhibited the protective effects of the innocuous level of lung thrombosis and nanoparticle-targeted endothelial delivery of the *Alox15* gene inhibited the severe thrombosis-induced increases in EC death and ALI in septic adult WT mice. These EC-targeted gene therapy-like approaches avoided the potential cofounding effects of genetic compensation in transgenic/knockout mice as used in the aforementioned studies and also modulate Alox15 expression only in ECs. Our findings together provide strong support for a pro-survival role of endothelial Alox15 in lung ECs during inflammatory lung injury induced by sepsis.

Finally, we employed a genetic mouse model of thrombocytopenia to show that platelet depletion diminishes sepsis-induced lung thrombosis and increases inflammatory lung injury and mortality in mice, and that the restoration of innocuous lung thrombosis protects against sepsis-induced inflammatory lung injury in the setting of thrombocytopenia. The inducible genetic model of thrombocytopenia enabled us to attenuate platelet levels and lung thrombosis without platelet-depleting antibodies or anti-coagulants, which also influence inflammation. By combining genetic thrombocytopenia with MB-induced lung thrombosis, we were able to diminish then restore lung thrombosis in a quantitative and stepwise manner in platelet-depleted mice; this finding was not entirely surprising, given that thrombi still form even when platelet numbers are reduced. It was also unsurprising that a relatively higher dose of MBs (i.e., 2,000 MBs/mouse) was required to induce the optimal protective level of lung thrombosis in thrombocytopenic compared with WT mice (i.e., 1,000 MBs/mouse), given the established contribution of platelets to thrombus formation. These studies help us understand the role of varying levels of thrombosis in the pathogenesis of ALI and the failure of anti-coagulant therapies in clinical trials of sepsis/ARDS patients ^34–37^.

In summary, our studies provide unequivocal evidence that: (i) mild lung thrombosis inhibits sepsis-induced ALI via endothelial Alox15; (ii) severe lung thrombosis increases sepsis-induced ALI, which can be attenuated by EC overexpression of *Alox15*; (iii) normalization of lung thrombosis inhibits thrombocytopenia-induced increases in ALI; and (iv) ARDS patients have reduced levels of *ALOX15*^+^ lung ECs, which validates the clinical relevance of our findings. Our surprising finding that mild lung thrombosis is protective may explain the lack of efficacy of anti-coagulant therapies seen in clinical trials of sepsis/ARDS patients. Thus, tuning of lung thrombosis to a protective level, or activation of ALOX15 signaling (e.g., through EC-targeted nanoparticle delivery of endothelial ALOX15 gene), represent promising therapeutic strategies against ALI/ARDS caused by sepsis.

## Materials and Methods

### Mice

Animal care and study protocols were reviewed and approved by the Northwestern University Feinberg School of Medicine Institutional Animal Care and Use Committee. Male and female C57BL/6J mice (aged 12-16wks) were used throughout the studies. For studies of thrombocytopenia (i.e., platelet depletion), *Pf4*Cre mice (The Jackson Laboratories, USA) were bred with *DTR* mice (The Jackson Laboratories, USA) to generate *DTR^Pf4Cre^*mice (referred to herein as PF4 mice). In these studies, littermate *Pf4*Cre^-^ mice (*DTR^Pf4-^*) served as WT controls (referred to herein as WT mice). To induce genetic thrombocytopenia, *DTR^Pf4Cre^* or *DTR^Pf4-^* mice received diphtheria toxin (DT) (20ng/g/day, i.p., Sigma-Aldrich, USA) for 3days. Blood was collected by cardiac puncture into EDTA-coated tubes and complete blood cell counts were performed using an automatic hematology analyzer (Advia 120, Siemens, USA).

### Nanoparticle-targeted EC-specific gene knockout and overexpression in adult mice

EC-specific disruption of *Alox15*, or *Cd22* expression in adult mice were achieved using nanoparticle-mediated genome editing as described previously ^41^. In short, EndoNP1 nanoparticles (Mountview Therapeutics LLC, USA) were incubated at room temperature for 10mins with the all-in-one plasmid DNA expressing Cas9 under the control of the EC-specific *CDH5* promoter and gene-specific guide RNA driven by the *U6* promoter following the manufacturer’s instructions, then administered intravenously (retro-orbitally) to C57BL/6J WT adult mice (40µg plasmid DNA/mouse). Seven days later, the genome-edited mice were used for studies. Mice treated with nanoparticles plus plasmid DNA expressing scrambled RNA served as wild type (WT) controls.

EC-specific *Alox15* overexpression in adult mice was also achieved using nanoparticle-mediated gene transduction. Briefly, EndoNP1 nanoparticles were mixed with plasmid DNA expressing mouse *Alox15* driven by *CDH5* promoter or empty vector (control) for 10mins at room temperature and then administered to C57BL/6J WT adult mice retro-orbitally (20μg of plasmid DNA/mouse). Mice were sacrificed at 62hrs post-administration.

### Mouse model of endotoxemia

To induce endotoxemia, mice received LPS (Santa Cruz Biotechnology, Inc, USA) at a dose of 3mg/kg body weight for sub-lethal injury studies, and 4-6mg/kg for survival studies, i.p. as previously described ^29, 40^.

### Mouse model of polymicrobial sepsis

To induce polymicrobial sepsis, mice underwent CLP as previously described ^40^. Briefly, mice were anesthetized with inhaled isofluorane (2.5% mixed with room air). When the mouse failed to respond to paw pinch, buprenex (0.1 mg/kg) was administered subcutaneously prior to proper sterilization of the skin with providone iodine, and then a midline abdominal incision was made. The cecum was exposed and ligated with a 3-0 silk tie 1 cm from the tip and the cecal wall was perforated with a 20-gauge needle. Control (sham) mice underwent anesthesia, laparotomy, and wound closure but no cecal ligation and puncture. Immediately following the procedure, 500ul of warmed normal saline was administered subcutaneously. Within 5min following surgery, the mice were able to wake from anesthesia. The mice received a second dose of buprenex at 6-8h post-surgery subcutaneously.

### Mouse model of pulmonary thrombosis

To induce lung thrombosis, mice received polystyrene microbeads (MBs, 15µm diameter, Invitrogen, USA) i.v. as previously described ^39^.

### Lung histology

Formalin-fixed paraffin-embedded lung sections (5μm) were stained with hematoxylin and eosin (H&E) or martius scarlet blue (MSB) as previously described ^39, 50^. Imaging was performed using a brightfield microscope (Zeiss Axio, USA) with mounted camera and integrated imaging software (ZEN 2.6, USA). Incidence of alveolar edema and septal thickening were counted in a blinded manner in H&E-stained lung sections. Incidence of thrombosis and fibrin area were assessed in a blinded manner in MSB-stained lung sections as previously described ^39, 50^. For each mouse, the incidence of edema, septal thickening, and microvascular thrombosis was counted in 10 randomly selected 20x fields. Percentage fibrin coverage was also assessed in 10 randomly selected 20x fields using image analysis software (ImageJ, NIH, USA). Data are expressed as an average per field.

### Lung vascular permeability measurement

Lung vascular permeability was assessed by measurement of lung EBA flux as previously described ^40^. Briefly, EBA (20mg/kg) was injected retro-orbitally 30mins before lungs were perfused with PBS free of blood. Lungs were then collected, blotted dry, weighed, and homogenized in 0.5ml PBS and then incubated with 1ml formamide at 60°C for 18hrs. Lung homogenates were centrifuged at 21,000g for 10mins. Optical density of the supernatant was determined at 620nm and 740nm.

### Myeloperoxidase (MPO) activity assay

Lung MPO activity was assessed as previously described ^29, 40^. Briefly, PBS-perfused lungs were homogenized in 0.5ml of 50mM phosphate buffer. Homogenates were centrifuged at 21,000g for 15mins at 4°C. Thereafter, the pellets were resuspended in phosphate buffer with 0.5% hexadecyl trimethylammonium bromide and subjected to a freeze-thaw cycle. Subsequently, the homogenates were centrifuged at 21,000g for 15mins at 4°C. Following addition of 34μl of sample to 10μl of O-dianisidine dihydrochloride (16.7mg/ml) and 50μl of H2O2 (0.015% v/v) in 1ml of phosphate buffer, absorbance was measured at 460nm every 20secs for 3mins.

### TUNEL staining

Lung apoptosis was assessed by quantification of TUNEL-positive cells in lung cryo-sections (5μm) using an In Situ Cell Death Detection Kit (Roche, USA) according to manufacturer’s instructions as previously described ^29^. Lung ECs were co-stained with anti-CD31 antibody (1:40 dilution, Abcam, USA) and anti-vWF antibody (1:250 dilution, Sigma, USA) for 18hrs at 4°C. Sections were incubated with Texas Red-conjugated secondary antibody (1:300 dilution, Invitrogen, USA) for 1hr at room temperature. Nuclei were counterstained with DAPI. Imaging was performed using a confocal microscope (Zeiss LSM 880, USA) with mounted camera and integrated imaging software (ZEN 2.6, USA). For each mouse, the number of apoptotic ECs (CD31/vWF^+^ TUNEL^+^) and the number of apoptotic non-ECs (CD31/vWF^-^ TUNEL^+^) were counted in a blinded manner in 10 randomly selected 25x fields. Data are expressed as an average number per 1,000 nuclei.

### EC isolation

CD31^+^ ECs and CD31^-^ non-EC fractions were isolated using magnetic-activated cell sorting as previously described ^40^. Briefly, PBS-perfused lungs were cut into small pieces and incubated in 1mg/mL collagenase A (Roche, USA) for 1hr at 37°C in a shaking water bath (200rpm). The lung samples were then dissociated into a single-cell preparation using the gentleMACS Dissociator (Miltenyi Biotec, USA) and filtered through a 40μm nylon cell strainer. The single-cell preparation was then blocked with 20% fetal bovine serum for 30mins and incubated with anti-CD31 antibody (Abcam, USA) for 30mins at room temperature, followed by capture of CD31^+^ ECs with magnetic anti-rat IgG-conjugated Dynabeads (Invitrogen, USA) for 30mins. EC specificity was confirmed by quantitative RT-PCR (qRT-PCR) demonstration of EC marker enrichment versus whole lung samples.

### RNA sequencing analysis

RNA expression was assessed in whole lung tissues by RNA sequencing analysis (Novogene, USA). Briefly, lung samples were collected after perfusion free of blood and homogenized with Trizol (Invitrogen, USA) for RNA isolation. Isolated RNA was further purified with RNeasy minikit (Qiagen, USA) with DNase I digestion to remove genomic DNA. Purified RNA was sent to Novogene (USA) for RNA sequencing analysis.

### Molecular analyses

To validate RNA sequencing data, RNA was isolated as described above. Following reverse transcription, qPCR analysis was performed using the ABI ViiATM 7 real-time PCR system (Thermo Fisher Scientific) with SYBR Green master mix to quantify the expression of gene of interest. Mouse cyclophilin (*Cypa*) was used as housekeeping gene for normalization. Nucleotide sequences are given in **Table S1**.

For quantification of genome editing efficiency, we employed a qPCR method as described previously ^41^. Isolated lung ECs and non-ECs were lysed in proteinase K solution overnight at 60°C and genomic DNA was purified by standard pheno:chloroform extraction. qPCR analysis of the wild-type genomic DNA without Indels was performed on genomic DNA samples using the ABI ViiATM 7 real-time PCR system (Thermo Fisher Scientific) with SYBR Green master mix. Nme1 genomic DNA amplification was used as normalization. Nucleotide sequences are given in **Table S1**.

Protein levels of Alox15 were measured in isolated lung ECs and non-ECs by enzyme-linked immunosorbent assay according to manufacturer’s instructions (Abbexa, USA). Protein levels are expressed per mg soluble protein as measured by the Bradford Assay according to manufacturer’s instructions (Bio-Rad, USA).

### Identification of potent guide RNA for in vivo study

Potent guide RNAs were identified for *in vivo* studies as previously described ^41^. Briefly, 4 guide RNAs for each gene were designed using the Genscript guide RNA database and the correspondent DNA oligoes were subcloned to the CRISPR*^CAG^* plasmid DNA expressing antibiotic gene against puromycin. Hepa-1c1c7 cells (ATCC, USA) were cultured in DMEM plus 10% FBS, 100U/ml penicillin, and 100μg/ml streptomycin. At 50-70% cell density, the Hepa-1c1c7 cells in complete medium were transfected with the all-in-one CRISPR*^CAG^* plasmid DNA using EndoNP1 nanoparticles. The transfected cells were then treated with puromycin (500ng/ml) for selection. The selected cells were then collected for genomic DNA isolation by standard phenol:chloroform extraction. QPCR analysis was performed on genomic DNA samples using the ABI ViiATM 7 real-time PCR system (Thermo Fisher Scientific) with SYBR Green master mix. Nme1 genomic DNA amplification was used as normalization. Nucleotide sequences are given in **Table S1**.

### In situ RNA hybridization (RNAscope analysis)

Formalin-fixed paraffin-embedded lung sections from patients with ARDS or from normal control subjects (unused donor lungs) were subjected to *in situ* RNA hybridization assay (RNAscope assay) for *ALOX15* mRNA expression according to manufacturer’s instructions (ACD Bio, USA). Lung ECs were co-immunostained with anti-CD31 antibody (1:40 dilution, Abcam, USA) and anti-vWF antibody (1:250 dilution, Sigma, USA) for 18hrs at 4°C. Sections were incubated with Texas Red-conjugated secondary antibody (1:300 dilution, Invitrogen, USA) for 1hr at room temperature. Nuclei were counterstained with DAPI. Imaging was performed using a confocal microscope (Zeiss LSM 880, USA) with mounted camera and integrated imaging software (ZEN 2.6, USA). For each individual, the number of ALOX15^+^ CD31/vWF^+^ ECs were counted in a blinded manner in 15 randomly selected blood vessels. Data are expressed as an average percentage of blood vessel ECs.

### Human lung samples

Archived post-mortem human lung samples from ARDS patients were obtained from the University of Illinois at Chicago Biorepository. Lung samples from unused donor lungs from the Pulmonary Hypertension Breakthrough Initiative (PHBI) were used as controls. The use of these archived samples was approved by the Ann & Robert H. Lurie Children’s Hospital of Chicago Institutional Review Board.

### Statistical analysis

Statistical analyses were performed using Prism 9 (Graphpad Software, Inc., La Jolla, USA). Data normality was assessed using Shipiro-Wilk tests. Two group comparisons were assessed with unpaired 2-tailed t-tests. Multiple group comparisons were assessed using one- or two-way ANOVA with Bonferroni post-tests. Differences in survival were assessed using the log-rank (Mantel-Cox) test. P values of <0.05 were considered significant.

## Data Availability

All data are presented and included in the manuscript.

## Acknowledgments

We are grateful to the Biorepository at the University of Illinois at Chicago for the sections of lung tissues from ARDS patients and to the Pulmonary Hypertension Breakthrough Initiatives (PHBI) for the lung sections from unused donor lungs. This work was supported in part by NIH grants R01HL123957, R01HL133951, R01HL148810, R01HL162299, and R01HL164014 to Y.Y.Z and by an AHA Career Development Award 19CDA34500000 to C.E.E.

## Author Contributions

C.E.E. and Y.Y.Z. conceived and designed the experiments. C.E.E., X.Z., and N.M. performed the experiments. C.E.E., X.Z., and D.W. analyzed the data. C.E.E., D.W., and Y.Y.Z. interpreted the data. C.E.E. wrote the manuscript. Y.Y.Z. supervised the project and revised the manuscript.

## Data and Materials availability

All data are presented and included in the manuscript.

